# Sex-specific DNA methylation in saliva from the multi-ethnic Fragile Families and Child Wellbeing Study

**DOI:** 10.1101/2022.12.22.22283872

**Authors:** Allison Reiner, Kelly M. Bakulski, Jonah D. Fisher, John F. Dou, Lisa Schneper, Colter Mitchell, Daniel A. Notterman, Matthew Zawistowski, Erin B. Ware

**Affiliations:** Department of Biostatistics and Center for Statistical Genetics, School of Public Health, University of Michigan, Ann Arbor, Michigan, USA; Department of Epidemiology, School of Public Health, University of Michigan, Ann Arbor, Michigan, USA; Survey Research Center, Institute for Social Research, University of Michigan, Ann Arbor, Michigan, USA; Department of Molecular Biology, Princeton University, Princeton, New Jersey, USA

**Keywords:** DNA methylation, sex differences, saliva, autosomal chromosomes, epigenetic epidemiology

## Abstract

The prevalence of many diseases differs by sex, potentially due to sex-specific patterns in DNA methylation. Autosomal sex-specific differences in DNA methylation have been observed in cord blood and placental tissue, but are not well studied in saliva or in diverse populations. We sought to characterize sex-specific DNA methylation on autosomal chromosomes in saliva samples from children in the Fragile Families and Child Wellbeing Study, a multi-ethnic prospective birth cohort containing an oversampling of Black, Hispanic and low-income families. DNA methylation from saliva samples were analyzed on 796 children at both ages 9 and 15 with DNA methylation measured using the Illumina HumanMethylation 450k array. An epigenome-wide association analysis of the age 9 samples identified 8,430 sex-differentiated autosomal DNA methylation sites at age 9 (P < 2.4×10^−7^), of which 76.2% had higher DNA methylation in female children. The strongest sex-difference was in the cg26921482 probe, in the *AMDHD2* gene, with 30.6% higher DNA methylation in female compared to male children (P < 1×10^−300^). Treating the age 15 samples as an internal replication set, we observed highly consistent results between the age 9 and age 15 measurements, indicating stable and replicable sex-differentiation. Further, we directly compared our results to previously published DNA methylation sex differences in both cord blood and saliva and again found strong consistency. Our findings support widespread and robust sex-differential DNA methylation across age, human tissues, and populations. These findings help inform our understanding of potential biological processes contributing to sex differences in human physiology and disease.

## Introduction

Health and disease outcomes differ between male and female children. For example, infectious diseases tend to show greater severity in male children than female children, purportedly due to the influence of sex hormones on immune function (1). Neuropsychiatric disorders, including autism spectrum disorder, bipolar disorder, and schizophrenia show differential prevalence by sex, a phenomenon linked to brain development differences in youth and adolescence (2, 3). Sex differences are largely attributed to the genetic contribution of the sex chromosomes, but are also influenced by sex hormones (4), differential gene expression (5), metabolites (6), and epigenetic patterns, including DNA methylation (4). Widespread differences in autosomal gene expression between sexes are noted across numerous human tissues, although the effect sizes are small (7).

DNA methylation (DNAm), the addition of a methyl group onto the fifth carbon of a cytosine residue in DNA, is itself associated with modulation of gene expression (8). DNAm is involved in genomic imprinting, in which genes are expressed in a parent-dependent manner, and in X chromosome inactivation, the silencing of gene expression on the X chromosome in female mammalian cells to achieve dosage compensation (8). Many DNAm sites remain stable over the life course and many other sites may change with development, age, and environmental inputs (9). Thus, DNAm can be an informative biomarker for disease, developmental stage, and environmental exposures (10) and may play a crucial role in sex-specific health outcomes in children.

Sex differences in DNAm are expected on the X chromosome in humans due to X chromosome inactivation (Hall et al., 2014; Duncan et al., 2018). However, sex differences in DNAm have also been reported on autosomal chromosomes in numerous tissues including blood (7, 11-13), buccal cells (14, 15), the prefrontal cortex (16), and the placenta (17, 18). The number of differentially methylated sites ranged from a few hundred to nearly ten thousand in these studies, depending on the tissue, DNAm array, and available sample sizes. All tissues except for placenta had a larger proportion of sites with higher DNAm in female subjects compared to male subjects. A recent meta-analysis identified 31,727 autosomal DNAm sites that were differentially methylated by sex in cord blood tissue of newborns and subsequently replicated in peripheral blood tissue samples (13). Further investigation of sex-specific DNAm patterns across autosomes can elucidate the underlying genes and biological mechanisms that contribute to sex differences in disease and health outcomes.

Most prior studies of sex and DNAm were cross-sectional investigations of either newborns or adults of European descent. The methylation profiles of adults are subject to accumulated lifetime exposures, which can alter inherited DNAm states and confound sex-specific profiles. Studies confined to single ancestry groups potentially limit generalizability. Moreover, few studies have biological samples at multiple time points on the same individuals. Studies that have analyzed sex differences in DNAm over time are based on cord blood and peripheral blood samples and identified largely stable sex-specific DNAm patterns from birth to late adolescence (13, 19). Few epigenetic studies of sex difference have been conducted in children and adolescents using saliva (15, 20), a more easily collected tissue (21). Those that have been conducted, favor non-diverse, convenience samples. In a volunteer sample of saliva tissue from 118 children aged 9 −14, 5,273 sites were differentially methylated by sex (FDR < 0.05) using the EPIC BeadChip (15). However, larger and more diverse studies of autosomal DNAm in the saliva of children are warranted to gain biologic insights.

This paper aims to characterize autosomal DNAm sex differences in saliva in a large, population-based sample from the Fragile Families and Child Wellbeing Study (FFCW), a longitudinal cohort of racially diverse children born to unmarried parents across large cities in the United States (22). We analyzed DNAm data assayed on the Illumina HumanMethylation 450k BeadArray from saliva samples obtained at two distinct time points, ages 9 and 15, on the same set of children assayed at the same time. We tested for sex-specific differences across the genome at both ages, evaluated consistency across the time points, and compared with results from prior studies of sex-differential DNAm in children.

## Methods

### Fragile Families and Child Wellbeing (FFCW) Study

The FFCW Study is a cohort study of 4,898 children from twenty cities in the United States (22). FFCW was designed to investigate the environmental and social factors that shape the development of at-risk children and contains an oversampling of Black, Hispanic and low-income families (22). Study personnel obtained baseline information on child participants and parents and/or caregivers at the time of the child’s birth, between 1998 and 2000. Follow-up data was collected at key developmental stages: ages one, three, five, nine, and fifteen. Interviewers collected information on relationships, attitudes, behaviors, mental and physical health, clinical health, economic and employment status, neighborhood characteristics, and demographic variables of parents and/or caregivers and children at each time point. Further details on the study design can be found at www.fragilefamilies.princeton.edu/documentation.asp. At age 9 and age 15, the focal children were interviewed directly and saliva samples were taken from children whose primary caregivers provided informed consent. The original participant data collection was approved by the Princeton University Institutional Review Board and this secondary data analysis was approved by the University of Michigan Institutional Review Board (HUM00129826). For this analysis, we accessed survey data through the Fragile Families and Childhood Wellbeing website and we received access to the DNAm data through the study Principal Investigators. The data will be released in 2023 per the NIH data guidelines.

A total of 3,400 FFCW children had survey data available at age 9, and 2,881 of these children provided a saliva sample. The FFCW measured DNAm on 837 of these children at age 9. Of those with methylation data at age 9, 817 children had methylation data for their age 15 saliva sample. Of these 817 children, we excluded those with discordant sex and those missing data for variables of interest: sex, poverty ratio, mother’s self-reported race/ethnicity, mother’s education, mother’s health status, and mother’s smoking status – all at baseline – and child’s age in months and child’s BMI at age 9. We treated the age 9 DNAm data as a discovery dataset and the age 15 DNAm data as an internal replication dataset.

### Demographic Measurements

Numerous measurements were collected on Fragile Families children and caregivers. We selected the following variables measured at child’s birth for inclusion in this analysis: child’s biological sex (male or female) as reported by the mother; child’s birth city (Detroit/Toledo/Chicago vs. Other); the mother’s poverty status, computed as the ratio of the mother’s reported household income to the United States Census Bureau national poverty threshold for the prior year; self-reported race/ethnicity (White non-Hispanic, Black non-Hispanic, Hispanic, or Other) and education level (less than high school, high school or equivalent, some college or technical school, or college/graduate school) of both the mother and father; maternal smoking habits during pregnancy (none, less than 1 pack per day, ≥1 packs per day), and self-reported maternal health status at baseline (within 48 hours of giving birth: great, very good, good, fair/poor). The BMI of the mother and focal child was collected at the home interview visits at age 9 and age 15. The child’s precise age in months was also recorded at the home interviews. Child’s birth city was dichotomized as Detroit/Toledo/Chicago vs. other because a subset of FFCW children were oversampled for DNAm measurements for use in the Study of Adolescent Neurodevelopment (23, 24), which investigated neurodevelopment among children primarily from Detroit, Toledo, and Chicago.

### DNA Methylation Data

Saliva samples were collected at ages 9 and 15 using the Oragene® DNA Self-Collection Kit (DNA Genotek Inc., Ontario, Canada) and shipped to Princeton University for extraction and processing. Samples from both time points were plated and processed simultaneously on the Illumina HumanMethylation450k BeadChip (Illumina, San Diego, CA). This array that contains probes for 485,512 DNAm sites across the genome (25). Plates contained a mix of age 9 and age 15 samples to mitigate potential batch effects.

DNAm image data was processed using the *minfi* package (26) and the *enmix* package (27) in R statistical software (version 3.5). The image data pairs (n = 1,811) were read into an RGChannelset using *minfi* and the *enmix preprocessENmix* function applied RELIC to correct for dye bias and out of band normalization to correct for background noise. The rcp function from *enmix* used applied linear regression calibration between correlated Type I and Type II probe pairs to adjust for probe type bias. For every sample, we measured the DNAm level at each site across the epigenome via a beta (β) value: the ratio of methylated fluorescent signal to total fluorescent signal (methylated + unmethylated signal) (28). The β-value is a continuous measure between 0 and 1 and is interpreted as the proportion of DNA copies that are methylated at a given locus for an individual. A value of 0 indicates all DNA copies are unmethylated at a given site, and a value of 1 denotes all DNA copies are methylated at that site (29). We define the beta matrix as the set of β-values across all probes for all samples.

We performed probe-level quality control filtering on the beta matrix, including both the age 9 and age 15 methylation data, and was visualized using a flow chart. We removed probes with a detection p-value > 0.01 or methylated/unmethylated bead count < 4 in more than 5% of samples (n = 47,930 probes) using *ewastools* (30). We then removed the remaining SNP probes (n = 59) and cross-reactive probes (n = 27,141 probes), identified via a Basic Local Alignment Search Tool (BLAST) search based on a list of known cross-reactive probes (31). N = 410,447 probes remained after filtering. We identified gap probes (outCutoff=0.01, threshold=0.05) with multi-modal distributions and probes mapping to sex chromosomes using the *minfi* package in Bioconductor (26). We annotated DNAm sites to genomic features using the Islands UCSC dataset from the *IlluminaHumanMethylation450kanno*.*ilmn12*.*hg19* package in Bioconductor (32). The hg19 genome build was used for gene annotation.

Individual children were filtered out based on the following criteria: >10% of sites with a detection p-value > 0.01 or bead count < 4 after removing previously mentioned poor quality probes (n = 34), discordant mother-reported sex and DNA methylation-predicted sex (n = 11), and if two sequential samples from the same individual exhibited genetic discordance between visits (n = 27). We removed samples with outlier methylation values, identified using the *enmix QCinfo* function (n = 6). We also removed technical replicates (one each from 49 pairs, preferentially selecting the first run sample as the “original”). After individual-level filtering, there were N = 796 children left. A sample dropped due to quality control at age 9 also eliminated the age 15 sample from analysis and vice versa.

We estimated the cell type proportion in each saliva sample using the Houseman algorithm implemented in the *estimateLC* function in the *ewastools* package (30), using the children’s saliva reference panel (33). This method uses a reference DNAm database and employs linear constrained projection to infer the proportion of epithelial and immune cells in saliva tissue (34).

### Statistical Analysis

All analyses were performed using R statistical software (version 4.0.3). Code to conduct analyses is available online (https://github.com/bakulskilab). We conducted analyses separately on the age 9 discovery data and the age 15 internal replication data. We calculated bivariate descriptive statistics and compared the distributions of demographic variables between the analytic samples (N = 796) and excluded samples (N = 2,604), and subsequently between male (N = 403) and female (N = 393) children in the analytic sample. We used Pearson’s chi-squared test for categorical variables and Wilcoxon rank-sum test for continuous variables using the *gtsummary* package (35) in R.

We conducted principal component (PC) analyses of the autosomal DNAm data. Variables with ANOVA association p-value < 0.05 for one of the top three principal components were considered potential confounding variables and included as covariates in regression modeling.

### Sex-specific Differential Methylation Analysis

We first tested for global DNAm differences by sex among autosomal sites. We computed a global methylation score per sample, defined as the average β-value per child across all probes. We tested for sex differences in the global methylation scores using a mixed effects model adjusting for potential confounding variables identified through the principal component analysis: main effect terms for epithelial cell proportion and mother’s race/ethnicity, and a random effect for plating batch.

We then performed an epigenome-wide association analysis (EWAS) to identify differences in individual methylation sites between male and female children. We excluded gap probes from this analysis. For each probe, we fit a linear model using the *lmFit* function in the *limma* package (36) with methylation β-value as the outcome and the sex of the child as the exposure of interest. We included fixed effect terms to control for epithelial cell proportion and mother’s self-reported race/ethnicity, and a random effect term for sample plate using the correlation argument in *lmFit*. The between-plate correlation was estimated using the *duplicateCorrelation* function in *limma*. The same model was used for both the age 9 discovery data and the age 15 replication data. The *lmFit* algorithm uses an empirical Bayes approach that computes a moderated t-statistic for each probe, for which the standard error is smoothed across all probes in the array for a more efficient standard error estimate (36). We used an epigenome-wide p-value significance threshold of 2.4×10^−7^ which is recommended for epigenome-wide association studies performed on the Illumina 450K array (37). As a secondary analysis, we ran the same main model using only DNAm sites located on the X chromosome.

### Sensitivity Analyses

We performed a sensitivity analysis on the age 9 sex-specific EWAS to confirm that the covariates included in our models properly accounted for latent sources of variation in the DNAm data. We estimated data-derived surrogate variables in the age 9 methylation beta matrix after protecting the effects of sex, epithelial cell proportion, and mother’s race/ethnicity using the *sva* function from the *sva* package (38). We added the resulting top ten surrogate variables to a model including sex, epithelial cell proportion, and mother’s race. We then compared the magnitude and significance of the sex regression parameters between the main model and the surrogate variable-adjusted model using Spearman’s correlation.

A second sensitivity analysis was performed to address potential effects of birth city on DNAm due to pollution and/or environmental differences. We repeated the EWAS analysis including a fixed effect term for SAND oversampled birth city (Detroit/Toledo/Chicago vs Other) in addition to the original list of potential confounding variables. We compared the sex-difference regression parameters to those from the main model.

### Differential Methylation Region Analysis

We used the *DMRcate* package (39) to identify differentially methylated regions between males and females. We used the recommended parameters lambda = 1,000 and C = 2, corresponding to a region being defined as a collection of at least two significant DNAm sites identified from the previous single-site analysis (P < 2.4×10^−7^) that were no more than 1,000 base pairs apart.

### Gene Set Enrichment Analysis

Gene ontology enrichment analysis was performed on sites that were significant in the age 9 sex-specific discovery analysis. We used the *gometh* function in the *missMethyl* package (40), which takes probe names and links them to DNAm sites on the 450K array and their corresponding Entrez gene IDs. The function employs Wallenius’ noncentral hypergeometric test and accounts for the uneven DNAm site distribution across genes. We report gene ontology terms overrepresented at FDR < 0.05.

### Genomic Feature Enrichment Analysis

We performed genomic feature enrichment analysis to determine the CpG island locations and regulatory elements that were enriched for differentially methylated sites identified at age 9. We mapped each DNAm site to North Shore, South Shore, North Shelf, South Shelf, CpG Island, or Open Sea regions using the Islands UCSC dataset from the *IlluminaHumanMethylation450kanno*.*ilmn12*.*hg19* package (32). We used a chi-square test of homogeneity to determine enrichment or underrepresentation of significant sites in each region, with the expected proportion per region equal to the proportion of all sites located in the given region. Enrichment analysis was performed for hypermethylated sites in males and hypermethylated sites in females individually, and as a whole. We next used the eFORGE tool (41) to determine enrichment of age 9 differentially methylated sites across fifteen different chromatin states from the Roadmap Epigenomics database, using recommended parameters of a 1,000 bp window size, a background repetition of 1,000, a strict p-value threshold of 0.01, and a marginal p-value threshold of 0.05. We tested the top 1,000 (the maximum allowable number of probes) most significant sites hypermethylated in males and females, separately.

### Internal Replication using Age 15 data

We performed an internal replication of the age 9 results using the age 15 data. We used the same procedures described for the age 9 data to detect gap probes. We performed a sex-specific methylation regression analysis across all autosomal sites passing QC in the age 15 data, employing the same main model. We tested each epigenome-wide significant age 9 site for replication in the age 15 data. We defined an age 9 site as replicated if the age 15 p-value for the site had a p-value smaller than a Bonferroni-corrected threshold accounting for the number of significant probes in the age 9 data and consistent direction of effect with the age 9 analysis. We also identified sites with sex-specific DNAm in the age 15 data using the epigenome-wide significance threshold of 2.4×10^−7^. We additionally conducted differential methylation region analysis and enrichment analyses for the age 15 data using the approaches described above.

### Comparison to Cord Blood Tissue

We compared our age 9 DNAm EWAS results in this FFCW analysis to a published set of differentially methylated sites from a meta-analysis in peripheral blood tissue in children aged 5.5 to 10 years across 9 cohorts from the Pregnancy and Childbirth Epigenetics consortium (13). Specifically, we used 40,219 DNAm sites identified as significant (P < 1.3×10^−7^) in a sample of N=8,438 newborns and replicated in an independent group of N=4,268 older children (P < 1.1×10^−6^). The main model was fit on methylation β-values and adjusted for sex, white blood cell proportion, and batch (13). We used Spearman’s correlation to compare adjusted effect estimates between the two studies.

### Comparison to Saliva Tissue in Prior Study

We compared our age 9 DNAm EWAS results in this FFCW analysis to a published set of differentially methylated sites identified in saliva samples of N = 118 children aged 9 to 14 years using the EPIC BeadChip (15). Participants were recruited from the San Francisco Bay Area, with 54% of the sample reporting as Caucasian. The main model adjusted for sex, age, ethnicity and estimated cell type proportion. We compared adjusted effect estimates for sex using Spearman’s correlation.

## Results

### Study Sample characteristics

The analytic cohort was composed of 796 children with both age 9 and age 15 methylation measurements that passed quality control, and non-missing data for covariates of interest (**Supplemental Figure 1**). We compared characteristics of the FFCW samples we included in our analysis relative to those we excluded in **Supplemental Table 1**. The analytic cohort was 50.6% male (n = 403) and 49.3% female (n = 393). The overall median poverty index was 1.4, with nearly 35% of the children born to families in poverty (poverty index < 1). The largest proportion (17.7%) of the cohort was born in Detroit, followed by Richmond, Austin, and Oakland (9.3%, 7.7%, 7.0%, respectively). In this sample, 55% of mothers self-reported as Black non-Hispanic; 21% of mothers were Hispanic, 20% identified as White non-Hispanic, and 3.4% reported as other. At baseline, 62% of mothers reported having a high school degree or less. Further, 80% of mothers reported no smoking behaviors at baseline. Male and female children had comparable values for poverty index, maternal self-reported race, maternal education level, and maternal smoking status at birth (**Table 1**). The median age 9 BMI was slightly higher in female children than male children, 18.4 compared to 17.8.

**Table 1.**
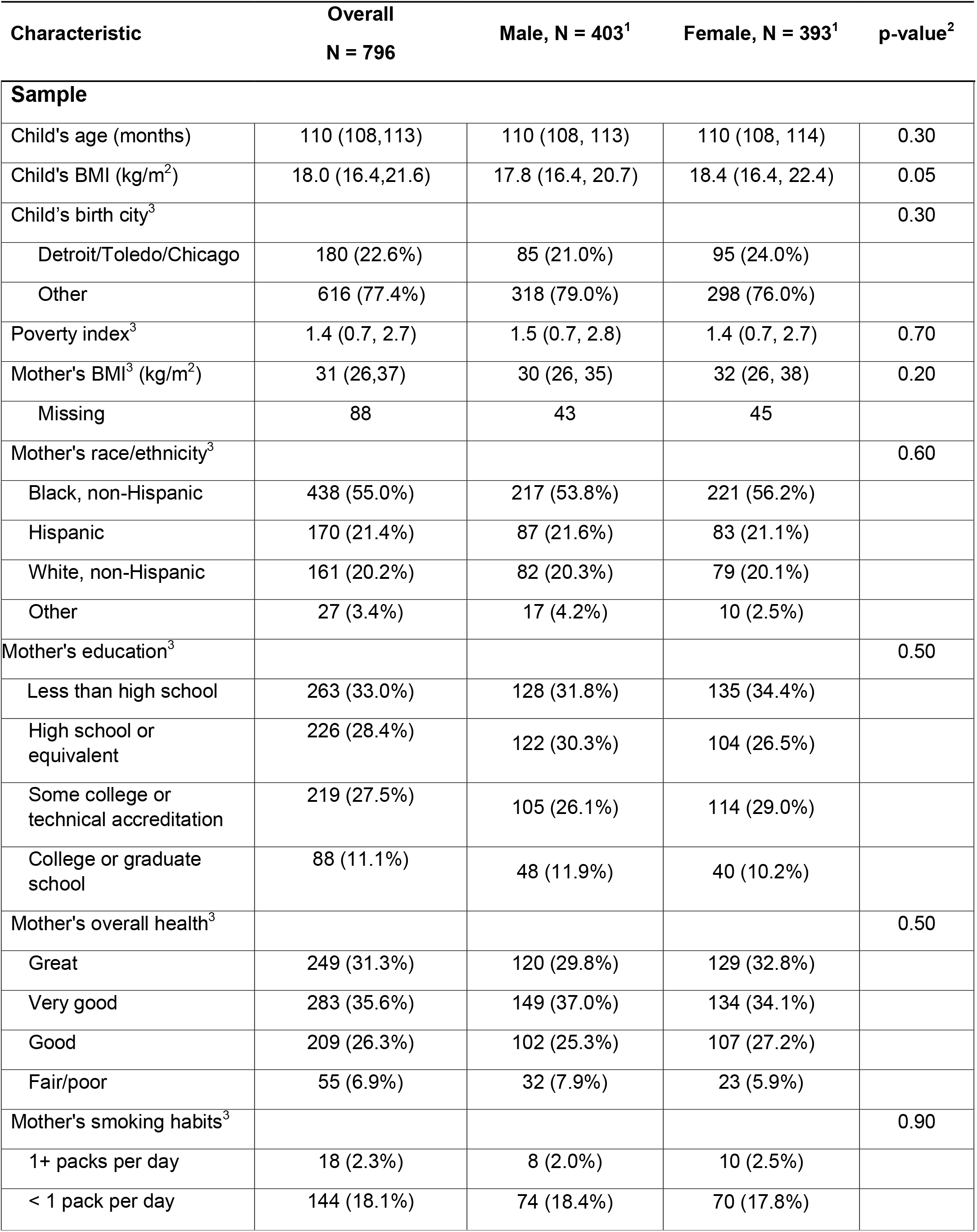
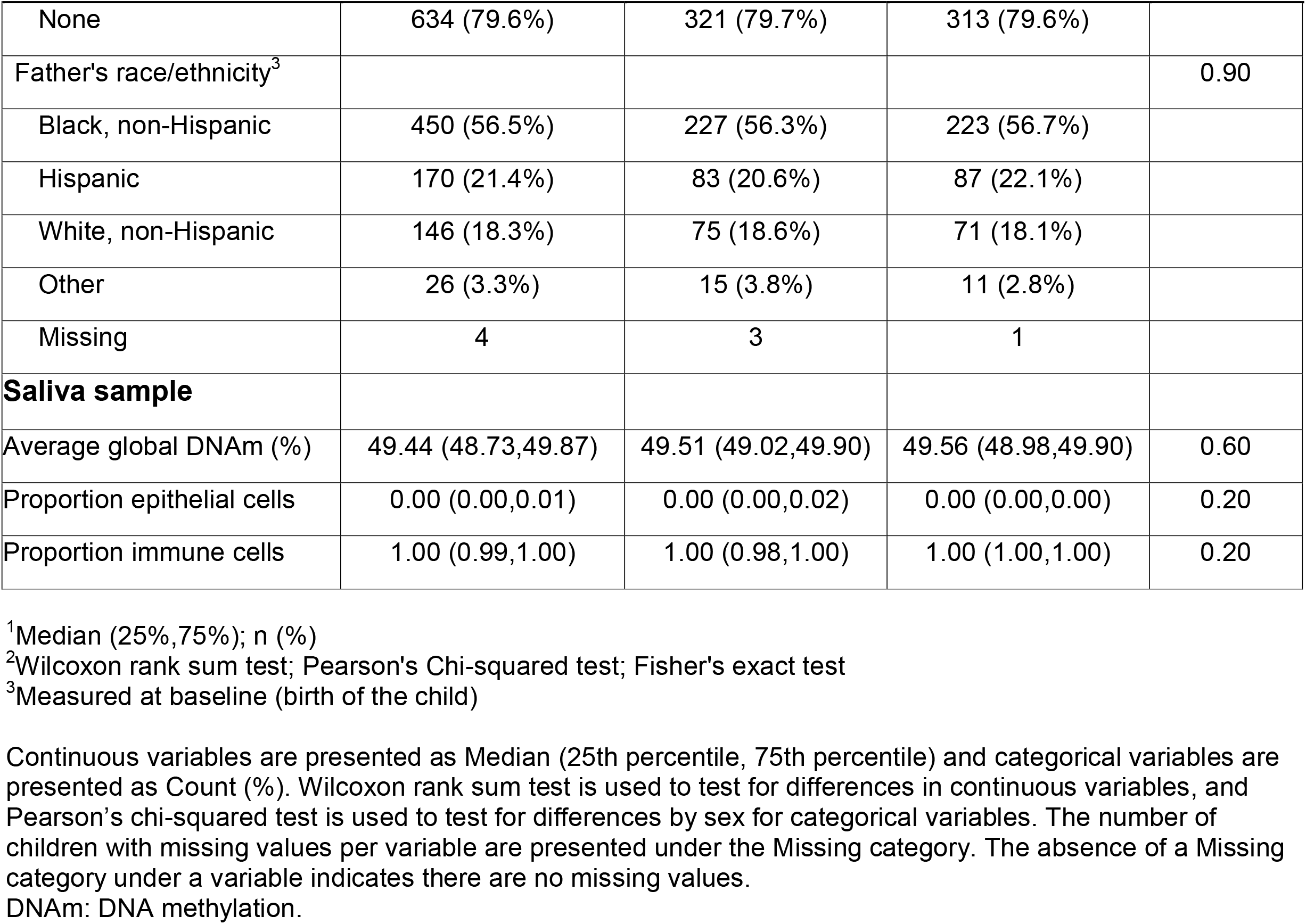
Fragile Families and Child Wellbeing Study sample characteristics at age 9, by sex (N = 796)

### DNA Methylation Data

We identified 410,447 DNAm sites that passed probe-level quality control filtering for both age 9 and age 15 measurements (**Supplemental Figure 2**), including 401,545 autosomal sites. We excluded n = 8,896 sex-chromosome specific sites from the main analysis. The distribution of average β-values across samples per site at age 9 was bimodal with ∼70% of sites having β-value ≤ 0.10 (10% DNAm) or ≥ 0.90 (90% DNAm). The peaks correspond to sites that are either unmethylated in all samples (β = 0) or methylated in all samples (β = 1) (**Supplemental Figure 3A**). The distribution of average β-values was similar in the age 15 methylation data (**Supplemental Figure 3A**). We identified 12,581 sites inferred to be gap probes in age 9 data and 11,485 sites in age 15 data (**Supplemental Figure 3B**), leaving a total of 391,980 sites for the age 9 analyses and 392,967 sites for the age 15 analyses. Principal component analysis of the methylation data revealed that cell type composition, sex, and sample plate were associated with individual PCs (**Supplemental Figure 4**), and were thus controlled for in subsequent sex-specific analyses. We also adjusted for mother’s self-reported race/ethnicity to account for race/ethnic and/or ancestry differences in DNAm patterns, which have been reported in the literature (42, 43).

### Sex-specific DNA Methylation Differences at Age 9

We first compared global β-values between male and female children. Overall, the median global β-value at age 9 was 49.44% (SD = 0.96%), with female children having slightly higher, though non-significant median values than male children (median β-value: 49.56% vs. 49.51%) (**Table 1**). After adjusting for cell type proportion, sample plate, and mother’s race/ethnicity, the mean global β-value was higher in female children (sex regression parameter = 2.7%, P = 0.48), indicating a slightly greater genome-wide methylation in the female children.

We next performed a probe-level analysis to identify individual sites with differential methylation between sexes, controlling for cell type proportion, batch and mother’s race/ethnicity. We identified 8,430 autosomal DNAm sites differentially methylated by sex at age 9 of the 391,980 sites tested (P < 2.4×10^−7^; **Figure 1A**), indicating widespread differential methylation by sex in saliva. The majority of the significant sites (n = 6,425, 76.22%) had higher average methylation in female children compared to male children, and an average adjusted absolute difference of 2.8%. Notably, the mean beta-values for significant sites have a roughly uniform distribution across the [0,1] interval, which differs from the bimodal pattern of mean beta-values observed across all sites(**Figure 1B**). Full results for each CpG site ranked by association p-value for age 9 can be seen in **Supplemental Table 2**. The two most significant sites, cg26921482 and cg11643285, were both hyper-methylated in female children and annotated to the *AMDHD2* and *RFTN1* genes, respectively. The third most significant site, cg02325951, was hypermethylated in males and mapped to the *FOXN3* gene. Additionally, we found that 91.6% (8,151 of 8,896) of X chromosome sites were differentially methylated by sex with an average adjusted absolute difference of 22.2%.

**Figure 1.**
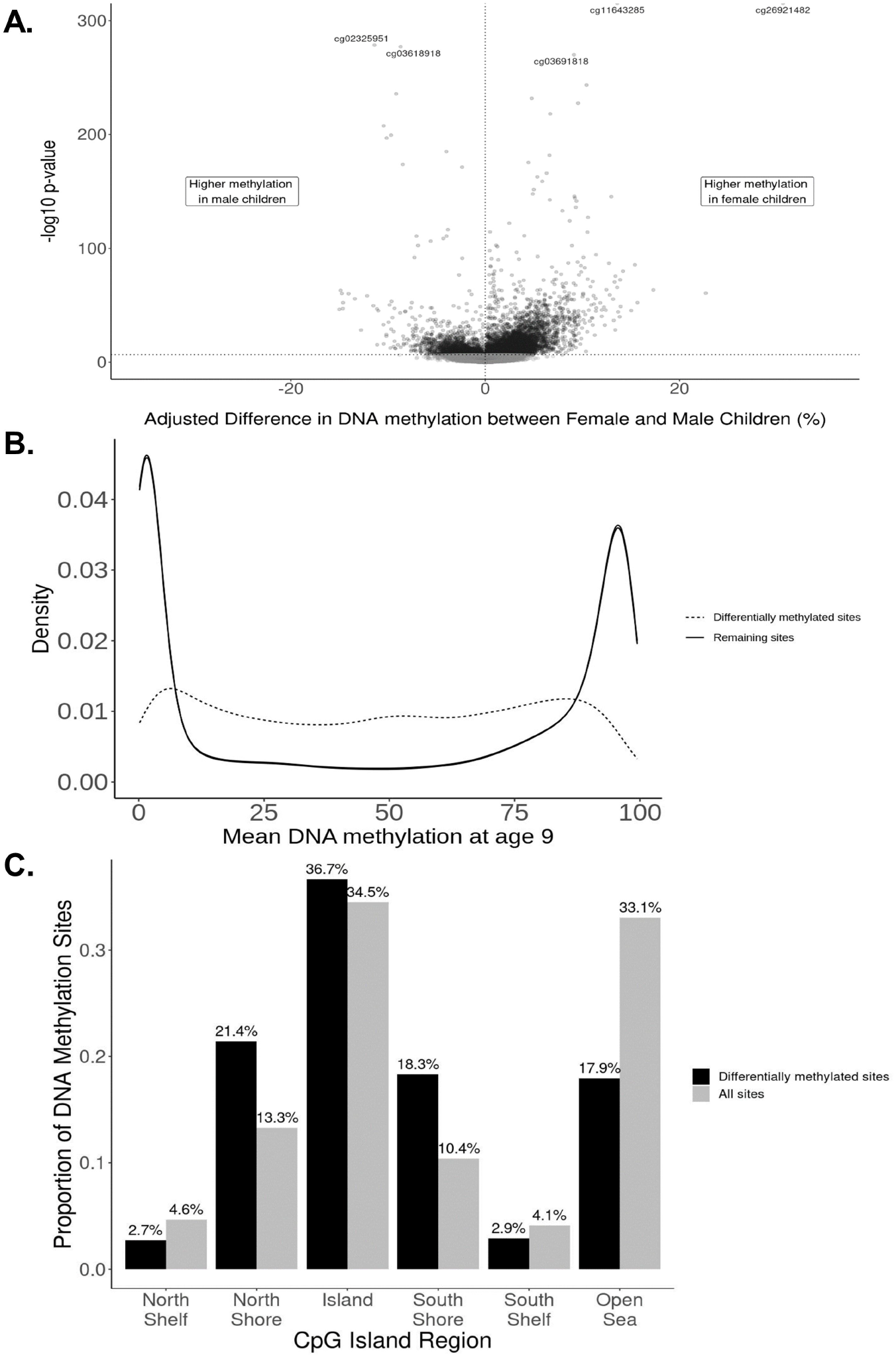
Characteristics of differentially methylated sites in the Fragile Families and Child Wellbeing Study sample, age 9 (N=796). **A:** Adjusted difference in autosomal DNA methylation between female and male children for each site (n=391,980 sites) in the Fragile Families and Child Wellbeing Study sample, age 9 (N=796). Effect sizes and –log10 p-values are from the main model containing fixed effects for sex, epithelial cell proportion, and mother’s race/ethnicity and a random effect for sample plate. Positive effect sizes indicate sites for which DNA methylation was higher in female children, while negative effect sizes represent sites for which DNA methylation was higher in male children at age 9. 8,430 sites achieved genome-wide significance at P = 2.4× 10^−7^ (horizontal dotted line), with n=6,425 (76.2%) having higher methylation in female participants. **B:** Distributions of mean DNAm for differentially methylated sites (n = 8,430; dashed line) and remaining sites (n = 383,550). **C:** Distribution of differentially methylated sites in relation to CpG islands. Fragile Families and Child Wellbeing Study sample, age 9 (N = 796). Proportion of significant sites (n = 8,430) from the main model that annotated to a given island region (black bar) are compared to the proportion of all sites (n = 391,980) annotated to that region (grey bar). Each regional comparison was significant at a Bonferroni-corrected threshold (α= 0.05/6 = 0.0083).

Our sensitivity analysis using surrogate variable adjustment produced p-values (r = 0.81) and sex-effect estimates (r = 0.93) strongly correlated with the main analysis, indicating that controlling for cell type composition, sample plate, and maternal race/ethnicity was likely sufficient. The sex regression parameter estimates were nearly identical (**Supplemental Figure 5**) and 83.4% of all significant sites were common to both models. Moreover, accounting for the birth city of the child resulted in little change in the sex-effect estimates (r = 0.99).

### Enrichment in Genomic Features among Differentially Methylated Probes at Age 9

The differentially methylated sites for which females had greater DNAm than males were enriched for biological processes related to behavior (P_FDR_ < 0.001), cell-cell signaling (P_FDR_ = 0.022), and regulation of ion transport (P_FDR_ = 0.041). These sites were also enriched in repressive polycomb regions (P_FDR_ < 0.01), which have been characterized by repressed gene expression (44). Conversely, sites with higher methylation in males were enriched in active transcriptional start sites (P_FDR_ < 0.01). None of the biological process gene ontology terms achieved FDR < 0.05 for sites hypermethylated in males. Differentially methylated sites were enriched within North Shore and South Shore regions, as well as CpG islands, but underrepresented in Shelves and Open Sea regions for both the female-hypermethylated sites and the male-hypermethylated sites (**Supplemental Table 3**). This pattern of enrichment remained the same for significant sites overall (**Figure 1C**).

### Differentially Methylated Regions at Age 9

We conducted differentially methylated region (DMR) analysis to identify gene-associated clusters of differentially methylated sites. We identified 1,499 DMRs between female and male children, with 1,197 regions (79.9%) characterized by higher average DNAm in female children. The regions of strongest significance were *HLA-DQB2, SCAND3, PPP1R3G* and *RP11-373N24*.*2* (top 15 regions presented in **Supplemental Table 4**). The DMR annotated to *PPP1R3G* was also identified in newborn cord blood tissue (13, 45). Among significant DMRs with the largest positive mean difference in sex were *PPFIA3* and *ZPBP2. PPFIA3* is involved in axon guidance and mammary gland development, while *ZPBP2* (Zona Pellucida-Binding Protein), expressed in the testis, is associated with sperm–oocyte binding during fertilization (46).

### Replication in Age 15 Methylation Data

We used the age 15 DNAm data as an internal replication set to confirm the strong sex-specific methylation in saliva at age 9. It is important here to reiterate that the age 9 and age 15 samples were plated and processed at the same time. Average β-values for individual sites were strongly correlated between age 9 and age 15 (Spearman Correlation = 0.9997; **Supplemental Figure 3A**). Gap probe identification was largely concordant (**Supplemental Figure 3B**) with 79.14% of gap probes identified at age 9 also flagged at age 15 and 86.69% of gap probes identified at age 15 flagged at age 9.

Of the 8,430 sites significant in the age 9 analysis, 7,421 (88%) were replicated among the age 15 DNAm data (Bonferroni alpha = 0.05 / 8430 = 5.93×10^−6^ and consistent direction of effect). Of the significant differentially methylated sites at age 9, ranked by p-value, the first 419 ranked sites were replicated at age 15 using the Bonferroni-corrected significance threshold. Of the 1,009 sites that did not replicate at this threshold, 99.8% had the same direction of effect in the age 9 and age 15 data. Ten sites were excluded during age 15 QC and not tested. Full results for each CpG site for age 15 can be seen in **Supplemental Table 2** (ranked by age 9 p-value).

We found that 64.31% of all significant DNAm sites were common to both time points at the epigenome-wide significance level, and had the same direction of effect and similar magnitudes (Spearman correlation = 0.99). The sex-difference effect sizes between the two time points were moderately correlated (r = 0.53; **Figure 2A**) across time points, while adjusting for cell type proportion, plating, and race/ethnicity differences. Thus, DNAm sites that were found significant at age 9 were likely to be significant at age 15. DNAm sites found significant at only one time point had p-values only marginally above the significance threshold at the other time point (**Figure 2B**). We identified 1,845 probes that were significant at the epigenome-wide significance level in the age 15 data that were not significant at age 9. Of these, 98.1% had the same direction of effect at age 9. Differentially methylated region analysis using the age 15 data identified 1,552 differentially methylated regions, resulting in a 71.46% concordance of region-associated overlapping genes across time points (**Supplementary Table 5**).

**Figure 2.**
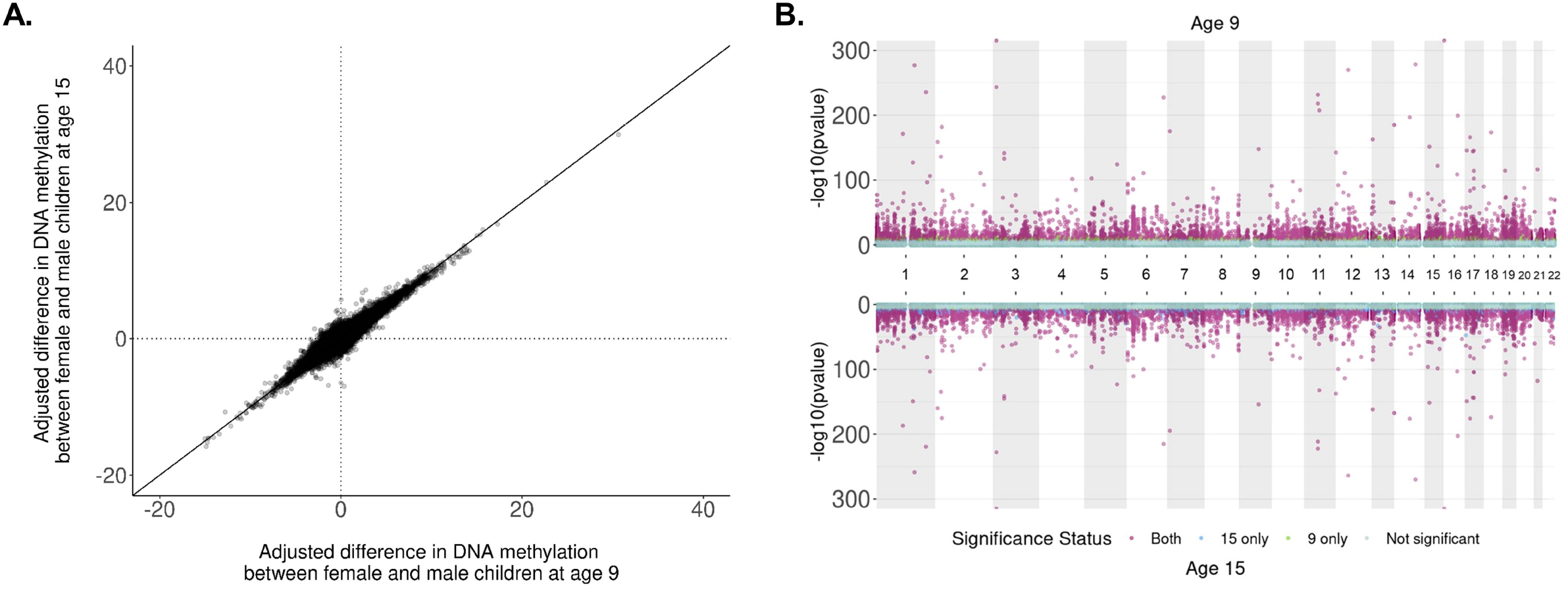
Comparison of adjusted sex differences in autosomal DNA methylation between ages 9 and 15 in the Fragile Families and Child Wellbeing Study (N=796, n = 390,659 sites). **A:** Correlation of adjusted difference in autosomal DNA methylation (%) by sex between ages 9 and 15 in the Fragile Families and Child Wellbeing Study (n = 390,659 sites). Effect sizes at each time point are from the main model adjusting for sex, epithelial cell proportion, mother’s race/ethnicity at baseline and a random effect for plate. Positive percentages are sites for which female children had higher DNA methylation than males. Spearman’s correlation = 0.53. **B:** Miami plot of sex-specific DNAm analyses at ages 9 and 15 in the Fragile Families and Child Wellbeing Study sample (N = 796). P-values are reported from the main model containing sex, epithelial cell proportion, maternal race/ethnicity at baseline, and a random effect for sample plate. A threshold of P = 2.4 × 10^−7^ is used for genome-wide significance.

### Comparison to Cord Blood Tissue

A previous study reported 40,219 sites differentially methylated by sex in peripheral blood tissue of children aged 5.5 to 10 years old at a p-value threshold of 1.1×10^−6^ (13). Of these sites, 37,413 were also tested in our analysis. Sites that were not tested (n = 2,806) were excluded as either a gap probe (n = 214) or due to quality control procedures (n = 2,592). We found a high correlation (Spearman correlation = 0.89) between sex-difference effect sizes from the age 9 analysis of saliva tissue and the peripheral blood tissue analysis (**Figure 3**) (13). Two DNAm sites were identified in the ten most significant sites in both analyses: cg26921482 on chromosome 16, *TBC1D24/AMDHD2;* cg17238319 on chromosome 3, *RFTN1*.

**Figure 3.**
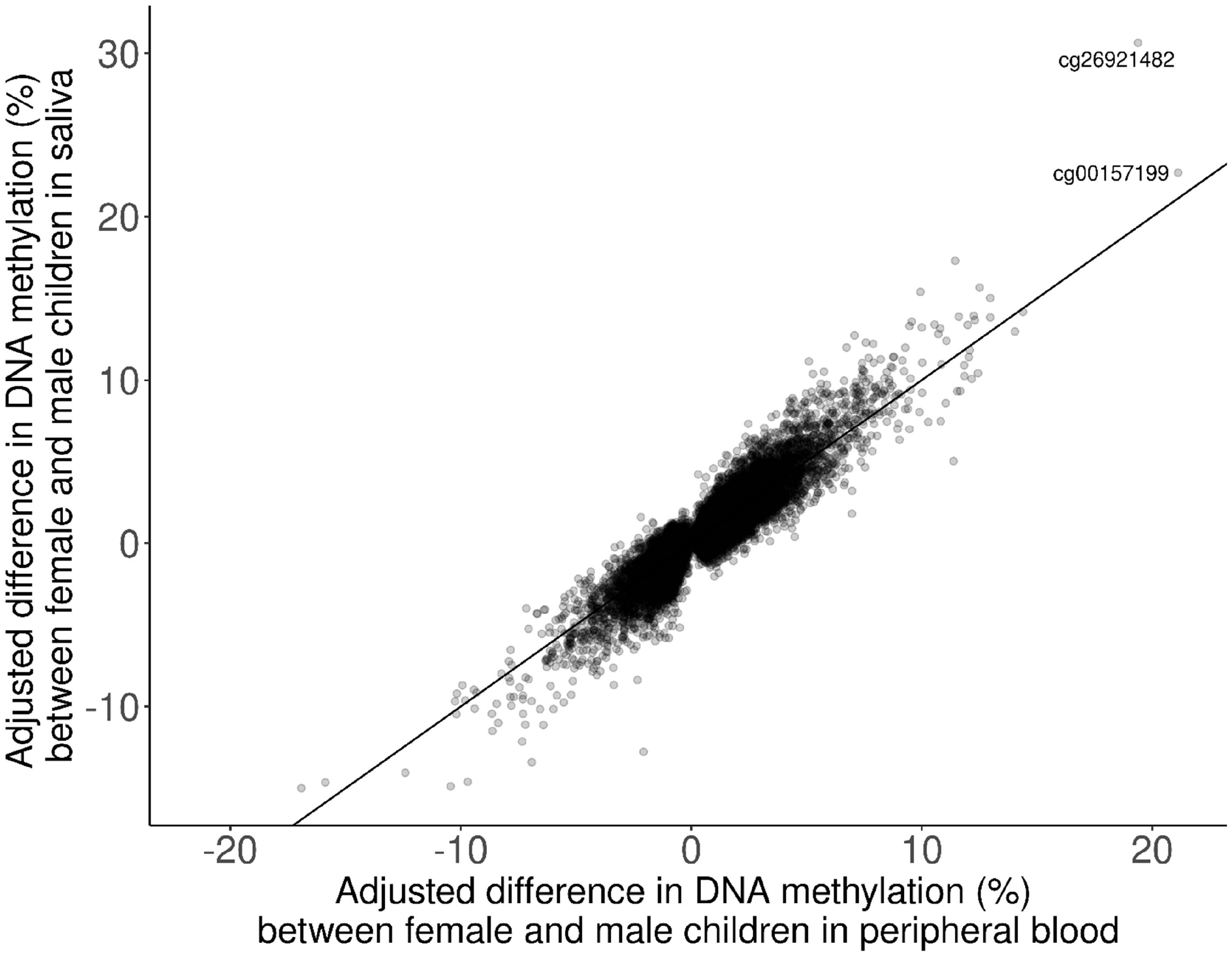
Comparison of adjusted differences in DNA methylation between saliva tissue from the Fragile Families cohort and cord blood tissue from Solomon et al., 2022 (Spearman correlation = 0.89)

### Comparison to Saliva Tissue

A recent study reported 5,273 sites differentially methylated by sex (FDR < 0.05) in saliva tissue of children aged 9-14 years (15). A total of n = 794,811 sites were tested using the EPIC BeadChip. Of the 5,273 significant sites, 1,885 sites were not tested in our analysis due to the use of different BeadChip technologies or removal during quality control. Of the 3,388 significant sites tested in both analyses, 2,476 sites were significant in both (73%). Of the 912 sites that were not significant in both analyses, 862 (94.5%) sites had the same direction of effect but magnitudes of effect were lowly correlated (Spearman correlation = 0.25). The effect estimates of the n = 3,388 commonly tested sites were moderately correlated (Spearman Correlation = 0.48).

## Discussion

In a large, diverse sample of children at ages 9 and 15 (n = 796), we observed widespread autosomal DNAm differences in saliva between male and female children. Specifically, we observed 8,430 sex-associated (P < 2.4×10^−7^) autosomal DNAm sites, and the majority (76.2%) of sites had higher DNAm in female children. Sex-specific DNAm sites were annotated to genes enriched for biologic pathways including behavior, cell signaling, and ion transport. These findings were consistent across DNAm measurement time points (Spearman correlation = 0.53) and consistent with prior literature in cord blood (Spearman correlation = 0.82) and saliva (Spearman correlation = 0.48). Together, these findings suggest that sex-specific differences in DNAm are present on autosomal chromosomes, and the findings are robust to tissue and time.

Our findings are in line with previous reports. For example, our most statistically significant DNAm site associated with sex was cg26921482, annotated to the Amidohydrolase Domain Containing 2 (*AMDHD2*) gene. In our study we found female participants had 30.6% higher DNAm. The Pregnancy And Childhood Epigenetics Consortium similarly observed that female infants had 23% higher DNAm at cg26921482 in cord blood at birth relative to male infants (13). Across these two studies in different tissues and developmental time periods, the DNAm differences observed are relatively large in magnitude (23-30%) and in a consistent direction. *AMDHD2* codes for a protein involved in amino-sugar metabolism and the hexosamine biosynthetic pathway, which is a minor branch of glycolysis (47). The hexosamine biosynthetic pathway may play a role in insulin resistance and diabetes (48). The Human Protein Atlas reports the AMDHD2 protein is present in higher levels in endocrine tissues and in male tissues including the testes (49). Similarly, the newborn Pregnancy and Childbirth Epigenetics consortium study observed a sex-specific differentially methylated position (cg11092486) annotated to the Protein Phosphatase 1 Regulatory Subunit 3G (*PPP1R3G*) gene with 15.2% lower DNAm in male participants relative to female (13). We also observed a sex-specific differentially methylated region associated with the *PPP1R3G* gene. The PPP1R3G protein plays a role in glycogen biosynthesis and lipid metabolism (50). DNAm differences at an *AMDHD2* site and in the *PPP1R3G* region by sex are consistent across tissues and developmental time, with implications for metabolism.

We observed that sex-specific DNAm sites were more likely to occur at the shores of CpG islands. For example, 10.4% of the DNAm array was annotated to the South Shore, while 18.3% of our sex-specific sites were annotated to the South Shore. Prior findings in pancreatic islet cells suggest that sex-specific differential DNAm mainly occurs at CpG shores, and not in CpG islands (51). Our sex-specific differentially methylated sites were enriched for repressive polycomb chromatin state regions, which are associated with repressed gene expression (44), and which play important roles in development and stem cells (52), as well as Alzheimer’s disease and cancer (53). Enrichment in repressive polycomb regions may suggest that these DNAm differences have implications for gene expression, and future studies may be able to link DNAm and RNA levels.

Many diseases and disorders have a sex-specific bias in risk or prevalence, and these same conditions have DNA methylation implicated in their pathophysiology. For example, schizophrenia is 1.4-times more likely to occur among males than females (54)). DNAm differences have been observed in brain tissue and blood comparing patients with schizophrenia to subjects without the disorder (55, 56). Furthermore, emerging evidence demonstrates the schizophrenia DNAm signatures may have sex-specific differences in DNAm (57). Similarly, most autoimmune disorders are more common in women, including lupus and rheumatoid arthritis (58). DNAm regulates immune cell differentiation, and dysregulation can induce immune cell auto reactivity, affecting the risk of autoimmune disorders (59).

Our study had several limitations, which may support future opportunities for research. Our study time points at children ages 9 and 15 likely spanned the pubertal window for many children (60). We were not able to include the timing of pubertal onset or duration in our models, and future studies may be able to track DNAm changes throughout puberty. We observed high correlation (Pearson correlation = 0.56) but not perfect correlation between sex-specific associations at the ages 9 and 15 time points, and other cohorts may be able to investigate longitudinal changes during this period in the context of puberty. Though we tested for replication with prior studies in other tissues, we were not able to identify many other cohorts with saliva DNAm for formal meta-analysis. The widespread findings observed in saliva in this cohort warrant a future meta-analysis and epigenetics consortia may be able to help facilitate these collaborations. We focused on autosomal chromosomes and provided results on sex chromosomes in the supplement to support further inquiries.

Because of our large sample size, our DNAm measures required several plates and thus potential technical artifacts resulting from batch effects. Consistent with other studies (61), we observed technical variation in the DNAm measures by sample plate and we adjusted for sample plate in our primary analyses. Our samples were randomized across plates by demographic factors including sex. To account for potential unmeasured confounding or technical variation, we additionally performed surrogate variable analysis with a sensitivity analysis, and observed our findings were robust.

Several factors contribute to the strength and breadth of this study. First, the study sample includes non-Hispanic Black and Hispanic participants who are currently underrepresented in genetic and epigenetic research (62). Ensuring the participation of diverse populations in research is important to assess generalizability of findings (62). Second, the study sample size of 796 is larger than many previous single cohort epigenome-wide association studies, which increases the study power to detect associations. Third, the study design includes repeated DNAm measures (from participant ages 9 and 15), which allowed us to assess persistence and reliability of measures. Importantly, samples from both ages were processed at the same time in the laboratory and participant paired samples from ages 9 and 15 were measured on the same slides (and thus plates), which minimized technical batch effects with respect to participant age. Fourth, we assessed participant sex using two methods (questionnaire and chromosome detection). Fifth, we detected DNAm in saliva, which is an emerging tissue type for epidemiologic research particularly in children because of the ease of collection. Sixth, we used a quantitative genome-wide array to detect DNAm that has been shown to have high reproducibility (63), and which is commonly used in epigenetic epidemiology (64) to promote replication. Seventh, we performed numerous sensitivity analyses, including surrogate variable analysis, pathway enrichment, and chromatin state enrichment to assess the robustness of the findings and increase the biologic interpretation of the findings. Eighth, to assess replication, we compared our findings to prior publications in cord blood and saliva (13). Together, the study population, design, and analytic approach are major strengths of this study.

In conclusion, we assessed autosomal sex-specific differential DNAm in children’s saliva at two time points in a large and diverse study population. We observed thousands of positions with differential DNAm, with predominantly higher DNAm in female samples. Our findings were also consistent with prior reports in other tissue types. Epigenetic epidemiology studies should take care to account for sex-specific DNAm patterns, even on autosomal chromosomes. Sex-specific DNAm positions were enriched for pathways of behavior and ion regulation, which may connect to different responses in these pathways by sex. Many diseases and disorders have prevalence differences by sex, and DNAm may be a marker or mediator linking sex and health.

## Data Availability

All survey data are available online through the Fragile Families and Childhood Wellbeing website and we received access to the DNAm data through the study Principal Investigators. The data will be released in 2023 per the NIH data guidelines.

https://fragilefamilies.princeton.edu/

## Acronyms

FFCWS: Fragile Families and Child Wellbeing Study
DNAm: DNA methylation
EWAS: Epigenome-wide association study
eFORGE: <u>e</u>xperimentally-derived <u>F</u>unctional element <u>O</u>verlap analysis of <u>R</u>e<u>G</u>ions from <u>E</u>WAS

## Funding

Training supported by: R25 AG053227 (AR, JF)

R01 MD011716 (CM, EW, JF)

R01 AG067592 (AR, KB, MZ, CM, EW, JF, JD)

R01 HD076592 (DN, LS)

## Acknowledgments

We thank the participants and staff of the Fragile Families and Child Wellbeing Study. We also wish to thank Brittni Delmaine for her editorial services.

## Author contributions

Allison Reiner: software, visualization, formal analysis, data curation, writing-original draft

Matthew Zawistowski: conceptualization, methodology, writing-review & editing, supervision, funding acquisition

Kelly M. Bakulski: conceptualization, methodology, supervision, project administration, writing-review & editing, funding acquisition

Erin B. Ware: conceptualization, methodology, supervision, project administration, writing-review & editing, funding acquisition

Jonah D. Fisher: conceptualization, validation, data curation

Colter Mitchell: validation, project administration, writing-review & editing

John F. Dou: validation, writing-review & editing

Lisa Schneper: investigation, writing-review & editing

Daniel A. Notterman: resources, project administration, investigation, writing-review & editing

