## Supplemental Figure for "Sex-specific DNA methylation in saliva from the multi-ethnic Fragile Families and Child Wellbeing Study"

**Affiliations:**

1415 Washington Heights

Ann Arbor, MI 48109

**Abstract (maximum word count: 250)**

The prevalence of many diseases differs by sex, potentially due to sex-specific patterns in DNA methylation. Autosomal sex-specific differences in DNA methylation have been observed in cord blood and placental tissue, but are not well studied in saliva or in diverse populations. We sought to characterize sex-specific DNA methylation on autosomal chromosomes in saliva samples from children in the Fragile Families and Child Wellbeing Study, a multi-ethnic prospective birth cohort containing an oversampling of Black, Hispanic and low-income families. DNA methylation from saliva samples were analyzed on 796 children at both ages 9 and 15 with DNA methylation measured using the Illumina HumanMethylation 450k array. An epigenome-wide association analysis of the age 9 samples identified 8,430 sex-differentiated autosomal DNA methylation sites at age 9 (P < 2.4x10^-7^), of which 76.2% had higher DNA methylation in female children. The strongest sex-difference was in the cg26921482 probe, in the *AMDHD2* gene, with 30.6% higher DNA methylation in female compared to male children (P < 1x10^-300^). Treating the age 15 samples as an internal replication set, we observed highly consistent results between the age 9 and age 15 measurements, indicating stable and replicable sex-differentiation. Further, we directly compared our results to previously published DNA methylation sex differences in both cord blood and saliva and again found strong consistency. Our findings support widespread and robust sex-differential DNA methylation across age, human tissues, and populations. These findings help inform our understanding of potential biological processes contributing to sex differences in human physiology and disease.

**Keywords:** DNA methylation, sex differences, saliva, autosomal chromosomes, epigenetic epidemiology

**Supplemental Figure 1**.

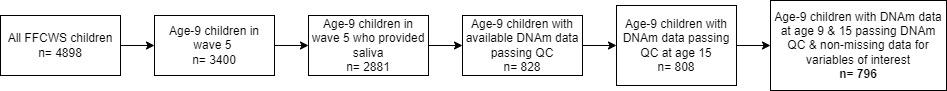

**Supplemental Figure 1**. Inclusion process flow chart for Fragile Families and Child Wellbeing Study analysis sample. Wave 5 refers to the data collection period when children were age 9. DNA methylation data is measured on the Illumina 450K BeadChip. Variables of interest include sex, poverty ratio, mother’s self-reported race/ethnicity, mother’s education, mother’s health status, and mother’s smoking status – all at baseline – and age in months and BMI at age 9.

FFCWS: Fragile Families and Child Wellbeing Study;

DNAm: DNA methylation;

QC: quality control.

**Supplemental Figure 2**

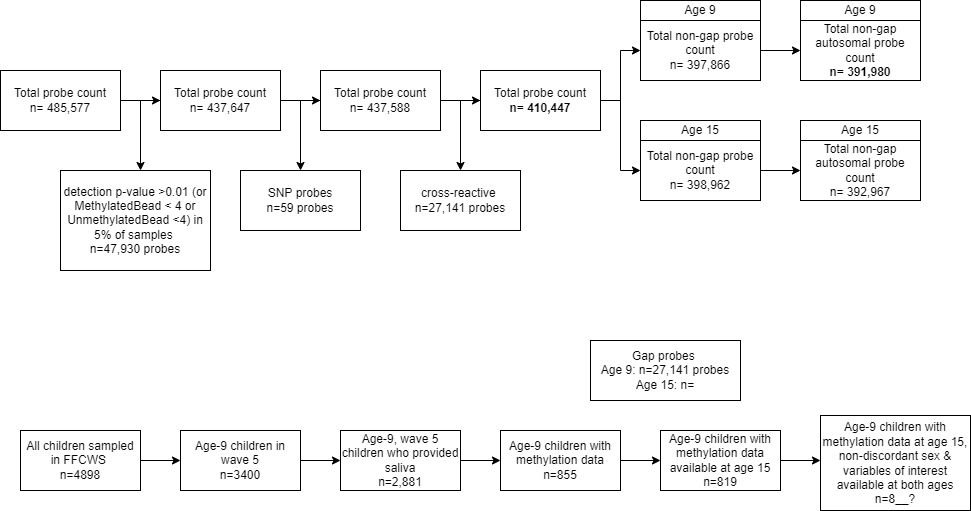

**Supplemental Figure 2**. DNA methylation probe quality control flow chart

SNP probes are those mapping to high frequency SNPs. Cross-reactive probes, which hybridize to homologous genomic locations to the intended target sequence, are obtained from a list by Chen et al. (2013). A total of n = 410,447 probes passed quality control at both age 9 and age 15. Sex-specific EWAS was performed on the age 9 methylation data excluding gap probes and sex chromosome probes (n = 391,980). SNP: single nucleotide polymorphism;

EWAS: epigenome-wide association study

**Supplemental Figure 3**

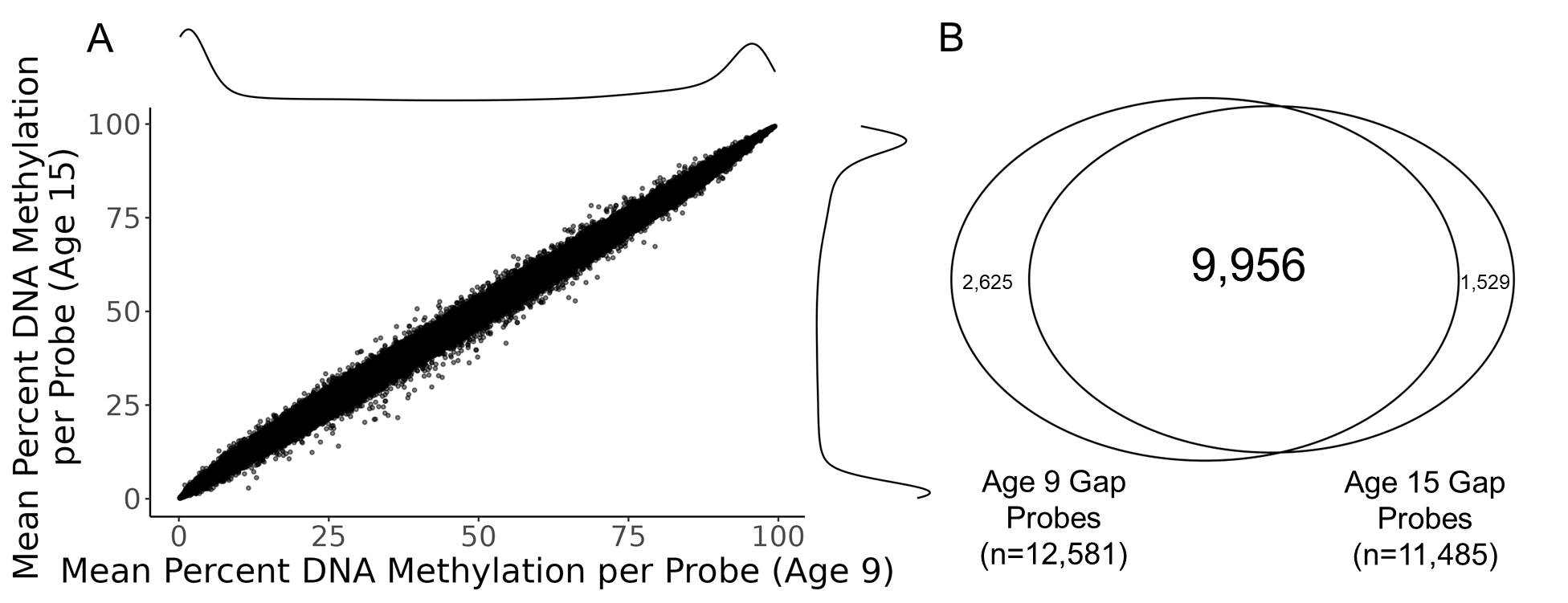

**Supplemental Figure 3:** (A) Comparison of average DNAm across children between age 9 and 15 measurements. The average β-value for each probe (n=410,447 probes) remains consistent between time points (Spearman Correlation = 0.9997). We note a bimodal density distribution at age 9 and age 15, with peak probe counts at β-values indicating either no DNA methylation (β = 0) or complete DNA methylation (β = 1). (B) Comparison of gap probe count between participants at age 9 and age 15. 9,956 probes were flagged as gap probes in the cohort at age 9 that were also identified as gap probes at age 15 (70.6% concordance). After removing gap probes, a total of 391,980 sites for the age 9 and 392,967 sites for the age 15 analyses remained.

**Supplemental Figure 4.**

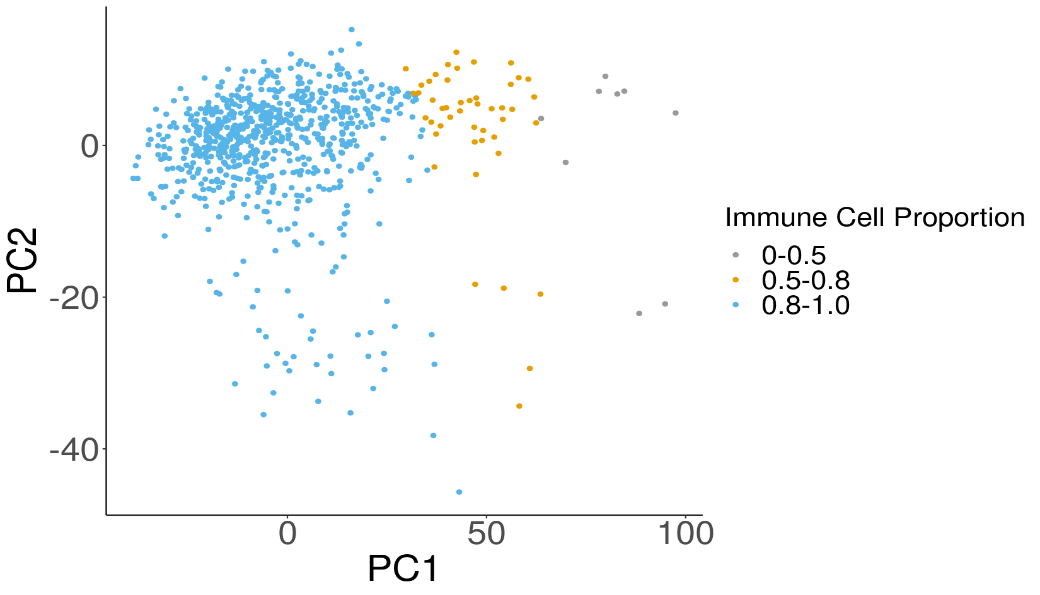

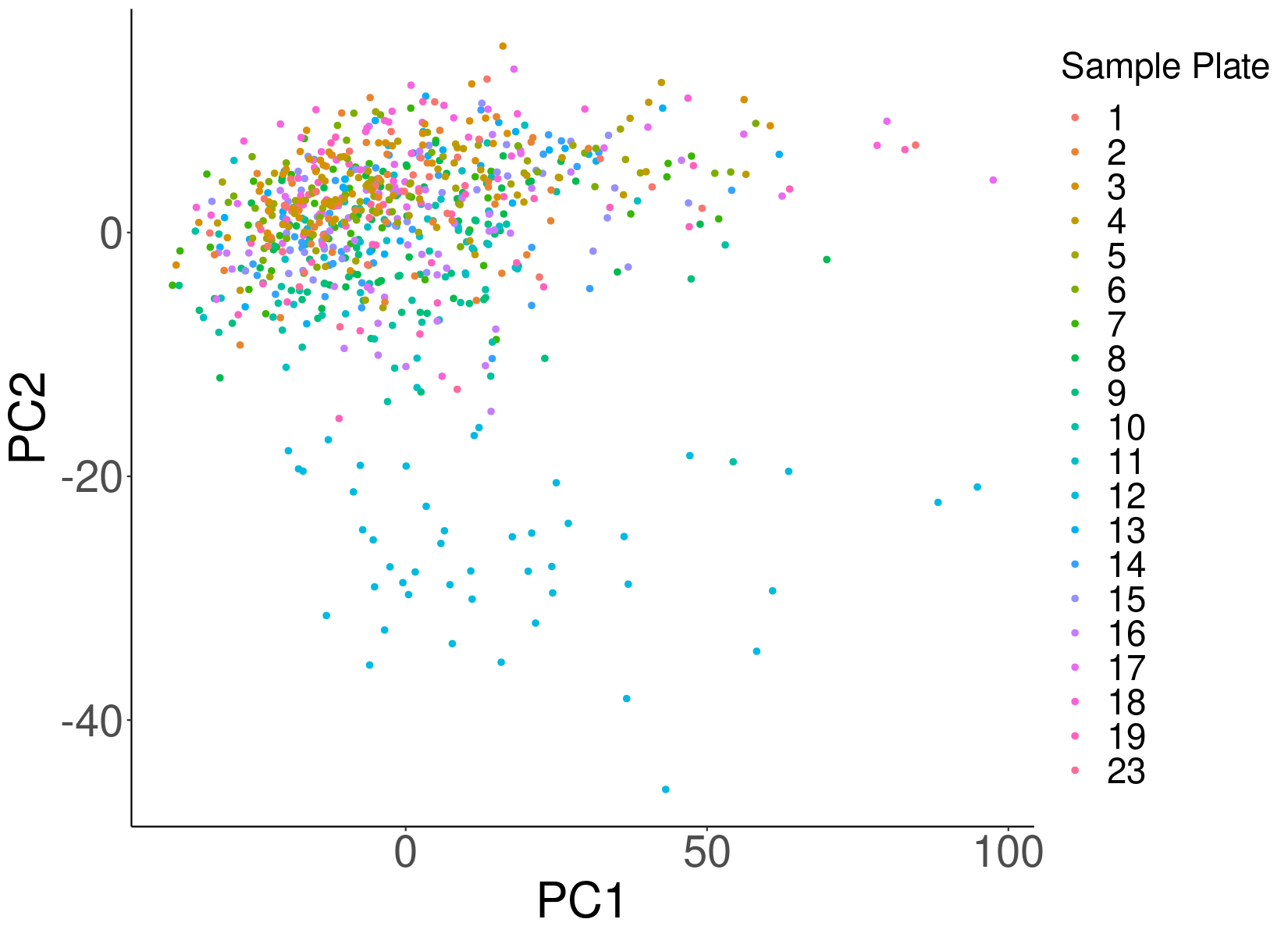

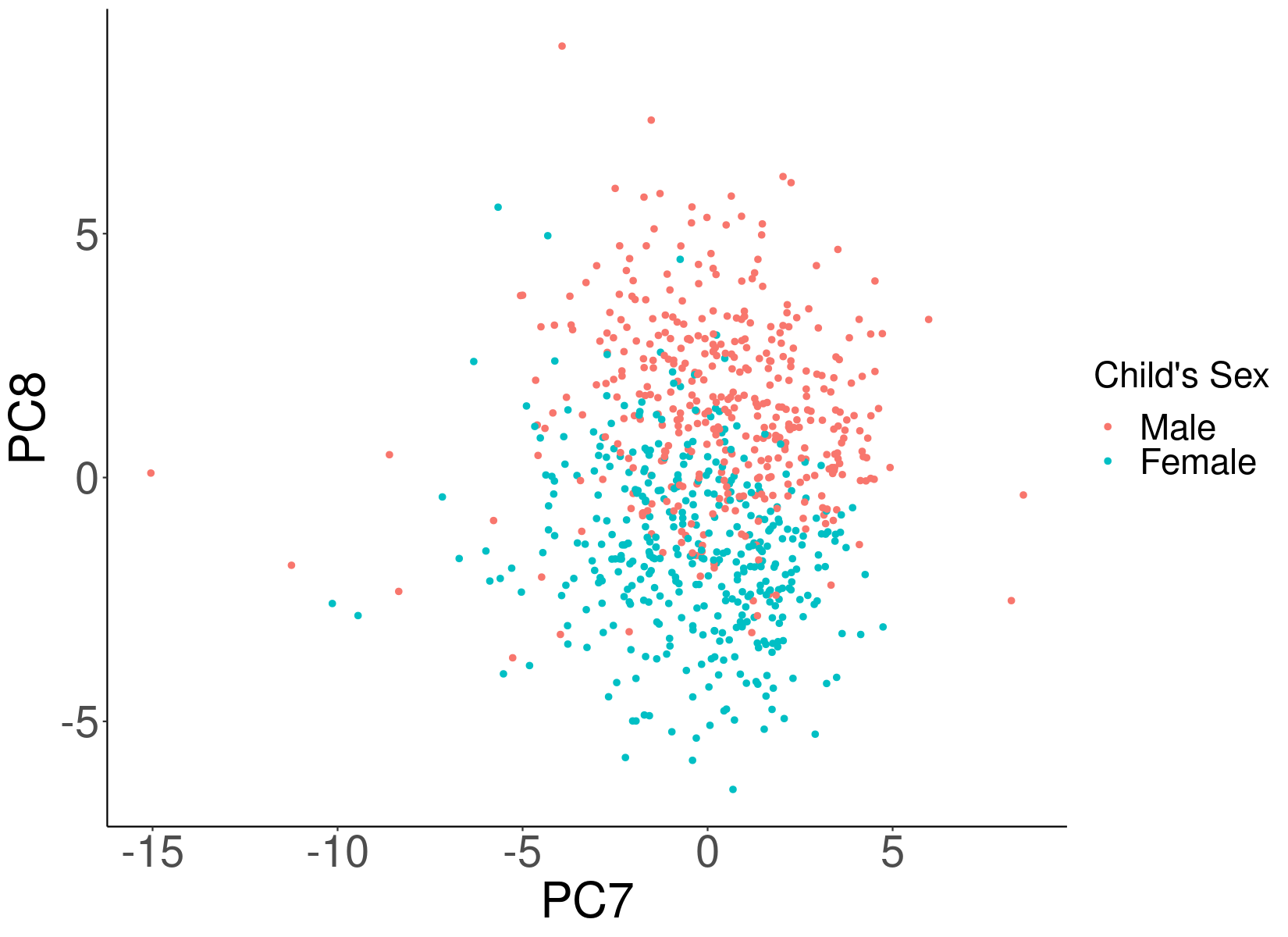

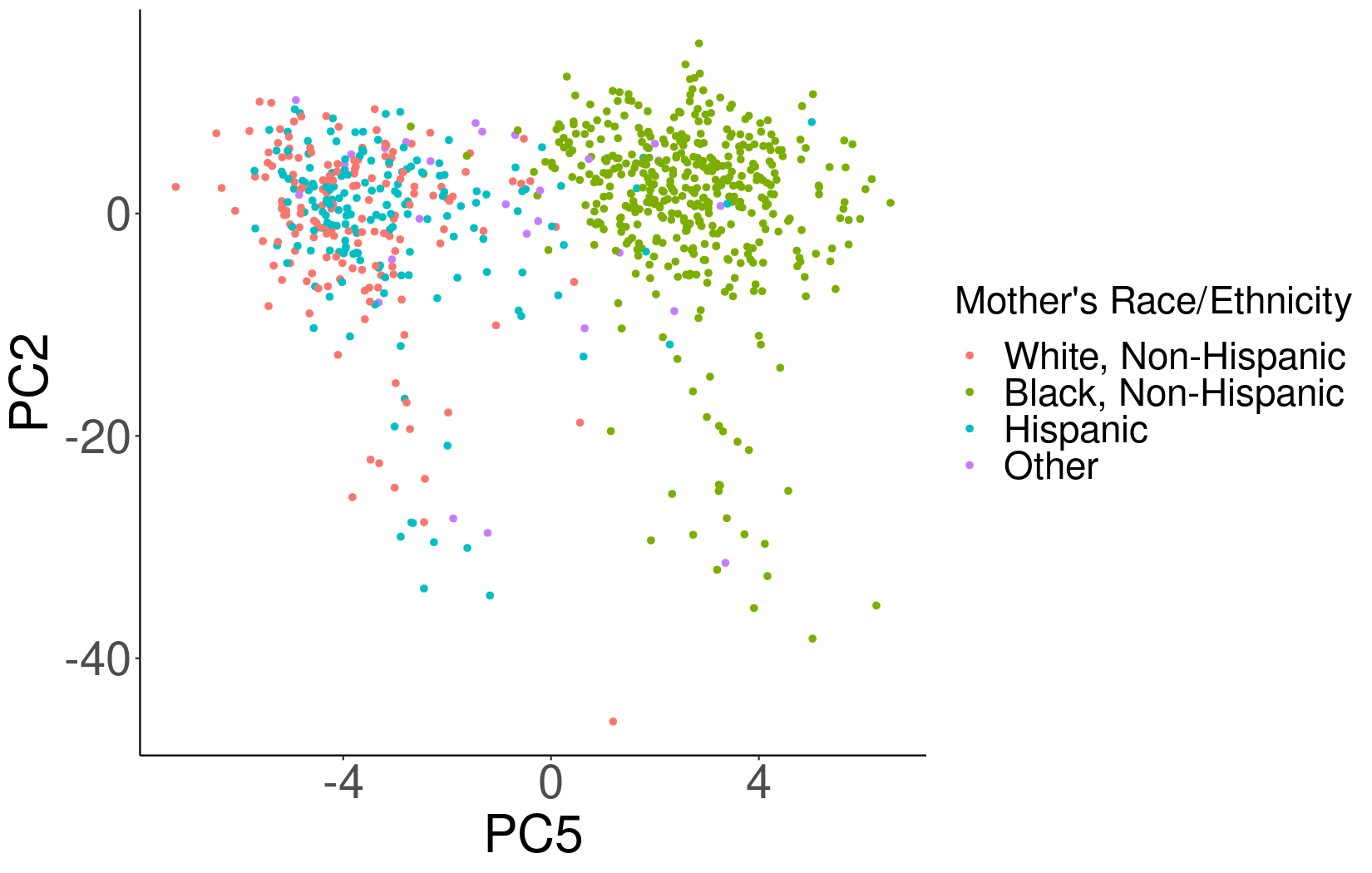

**A**

**B**

**C**

**D**

Supplemental Figure 4. Principal Component Analysis plots of age 9 DNA methylation data in the Fragile Families and Child Wellbeing Study, n = 391,980. Sample plate (A), Immune cell proportion (trichotomized) (B), and mother’s self-reported race/ethnicity (C) were associated with at least one of the top three principal components (p < 0.05). Child’s biological sex (D) was most strongly associated with PC7 (P = 1.39 x 10^-6^) and PC8 (P = 7.61 x 10^-89^). PC1 refers to the first principal component, explaining the most variation in the DNA methylation data. PC, Principal Component.

**Supplemental Figure 5.**

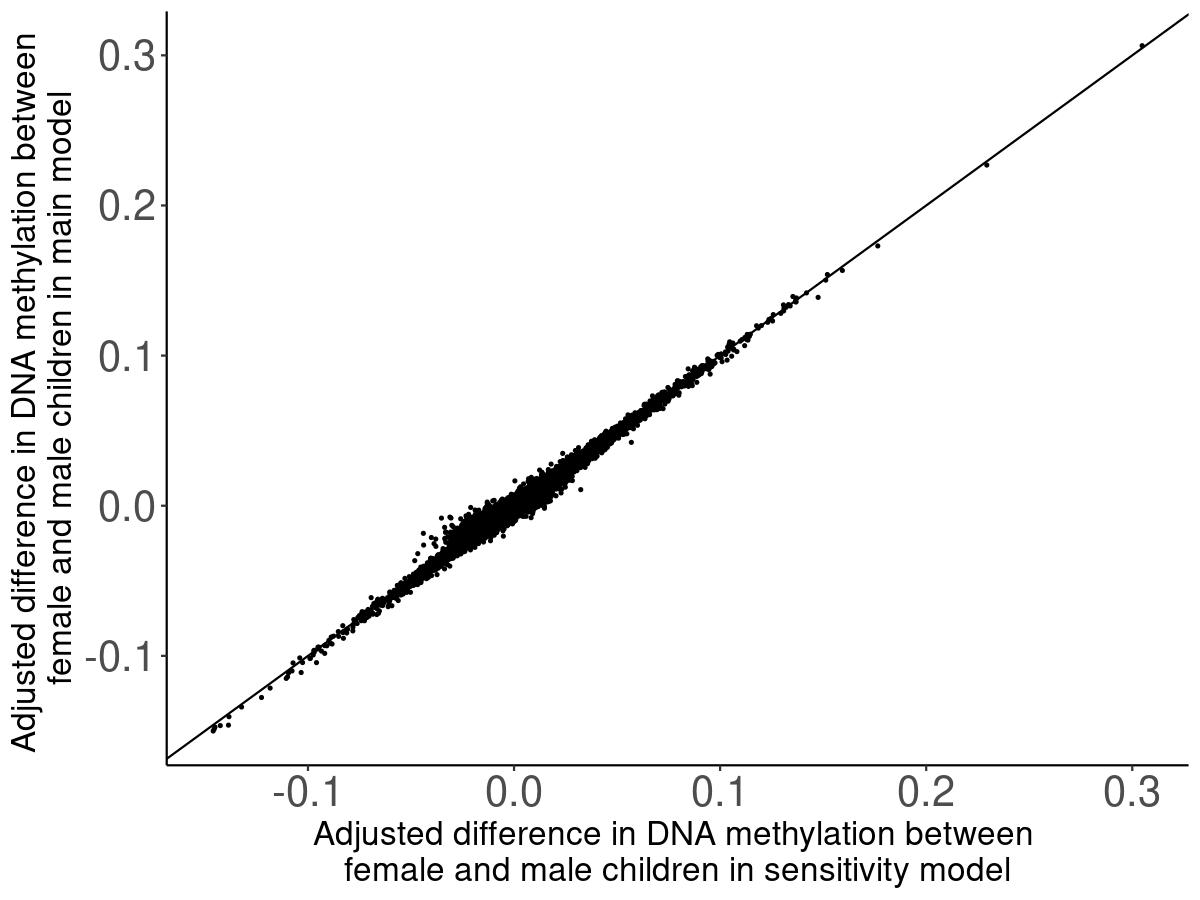

Supplemental Figure 5: Comparison of effect estimates from the main model to the surrogate-variable model (Spearman correlation = 0.93). The main model includes fixed effects for sex, epithelial cell proportion, mother’s race/ethnicity and a random effect for sample plate, and the surrogate-variable model contains fixed effects for sex, epithelial cell proportion, mother’s race/ethnicity and the top ten surrogate variables estimated from the age 9 DNAm data.
